# Supportive Strides: Co-designing Community-Based Walking Programs with Embedded Socialisation and Peer Support for People after Stroke

**DOI:** 10.64898/2026.09.17.26362966

**Authors:** Natalie A. Fini, Priyanka Cahill, Brian A. Beh, Louise Suttie, David Gallant, Andrew Martel, Iderlina Mateo-Babiano, Claudia H. Marck, Kathryn S. Hayward, Emily Ramage

## Abstract

**Purpose:** To co-design a community-based walking program that incorporates social connection to improve physical activity post-stroke.

**Methods:** A four-stage Integrated Knowledge Translation approach was used. Stage 1: research team (2 stroke survivors, 8 professionals) defined research questions, scope and plan. Stage 2: workshops held with stroke survivors, carers, and health and design professionals to identify important program principles. Stage 3: research team used a Nominal Group Technique to make key decisions and develop the program. Stage 4: program presented to participants; feedback gathered and adaptations made. Workshop findings were iteratively analysed and summarised during sessions.

**Results:** Twenty-six participants were recruited across stages 2 and 4 (12 stroke survivors, 2 carers, 12 professionals). Key program principles identified in Stage 2 were social support, inclusive environment, motivational support, co-ordination and administrative support. The Supportive Strides program was co-designed to incorporate these principles. Program elements included accessible outdoor walking routes, pre-walk coffee and social connection, peer-support, online group chat, milestone celebrations, and a safety checklist. Following Stage 4 participant feedback, adaptations were incorporated e.g., simpler screening process, include more walking route information.

**Conclusions:** The co-designed Supportive Strides community-based walking program is ready for testing to improve physical activity and social connection after stroke.

**Implications for Rehabilitation:**

- The Supportive Strides community-based walking program with embedded socialisation and peer support was co-designed by stroke survivors, carers, and health and design professionals
- Socialisation and peer support are considered critical elements for engagement and sustainability of post-stroke physical activity programs
- Inclusive environmental design may be important for outdoor walking activities for stroke survivors
- Tracking progress, celebrating achievements and coaching support are motivators for improving post-stroke physical activity levels

## Introduction

Being physically active is an imperative for living well after stroke. ^1^ Meeting physical activity guidelines can help to prevent recurrent stroke. ^2^ Yet, physical activity remains a challenge for people after stroke who are highly sedentary and inactive. Stroke survivors take less than half the number of steps per day compared to non-stroke age-matched adults^3^ and struggle to embed physical activity behaviours into their lives long term after stroke. ^4^

Poor physical activity can be compounded by social isolation and loneliness. It has been proposed that social networks and peer support can improve engagement in physical activity as well as functional recovery. ^5–8^ Evidence from healthy older adults and chronic disease populations demonstrates that including a social or peer support component in walking-based physical activity programs can improve uptake, engagement and physical activity behaviours long term. ^9–14^ These types of programs have not been tested in a stroke-specific population. In Australia, there are opportunities for social connection combined with group-based walking physical activity through the Heart Foundation. ^13^ They run a highly successful community-based walking program, “walking wins,” to improve health and physical activity. ^13^ It has substantial reach and retention in people with a range of health conditions (e.g., cardiovascular disease, arthritis). ^13^ However this program was not designed with the specific needs of stroke survivors in mind.

Due to the long-standing physical, communication, emotional and cognitive disabilities persisting after stroke, additional considerations and support are often required to participate in group-based initiatives. When stroke survivors do participate in physical activity, walking is the most common form of physical activity chosen. ^15^ There are many potential reasons for this – there is no associated cost, no requirement for complex equipment or a very high level of physical ability, it is safe, and it can be done in one’s local environment at a flexible pace or duration. Therefore, a walking group could be a way to engage stroke survivors in regular physical activity while providing social connection. Furthermore, people with stroke often like to exercise with others who have had a stroke. ^16^ The social component of a stroke walking group may help combat social isolation, loneliness, and facilitate physical activity engagement and sustainability.

When developing healthcare interventions, it is important to involve end-users in intervention co-design. ^17, 18^ Co-design is thought to increase relevance, practicality and uptake ^19^ and has been used successfully to develop several physical activity-based stroke recovery interventions. ^20–23^ Therefore, this study aimed to co-design a community-based walking program with embedded socialisation and peer support for people with stroke.

## Materials s Methods

### Study Design

This study used a four-stage Integrated Knowledge Translation approach to co-design a community-based walking program with embedded socialisation and peer support. The four stages are outlined below in the procedure section and figure 1. Core principles of the Integrated Knowledge Translation approach include partnership between researchers and people who will use the research, respect, feedback to improve the process and product, confidentiality, and use of common language. ^24^

**Figure 1:**
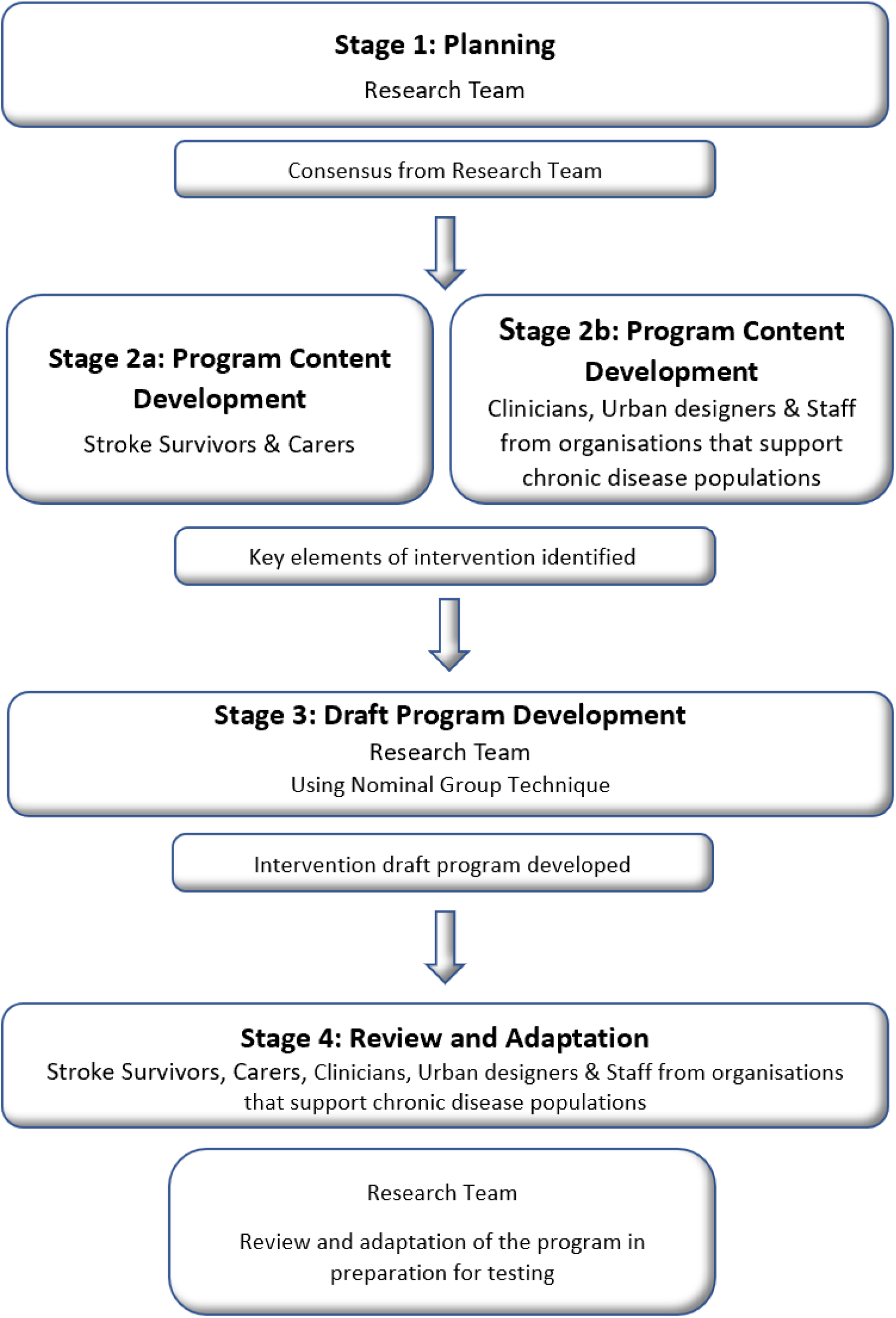
Four-Stage Integrated Knowledge Translation Approach (adapted from Ramage et al., 2022 to include participant groups specific to this project, Nominal Group Technique and final research team review in Stage 4). ^22^

Nominal Group Technique methodology^25^ facilitated consensus within the research team on important program principles in Stage 3. The Guidance for Reporting Involvement of Patients and the Public, Version 2 (GRIPP2) reporting guidelines^26^ were adhered to throughout this study to enhance transparency and quality of reporting (see appendix 1). Ethical approval was received from the University of Melbourne Human Ethics Committee (ID: 2025-32829-68681-3) and all participants provided informed consent.

### Contributors

#### Research Team

The research team refers to the named authors on the manuscript and includes researchers from a range of backgrounds (physiotherapy, sport development and management, architecture and urban design, and public health). Critically, the research team also included two people with lived experience of stroke with professional backgrounds in communications and nursing who were remunerated for their participation and input.

#### Study Participants (Knowledge-User Informants)

We aimed to recruit two groups of knowledge-users or study participants. The first group were people with lived experience of stroke – either stroke survivors or carers of stroke survivors. The second group were termed “professionals” and included: clinicians (health workers) of any discipline with experience in stroke; urban designers with expertise in disability access or walkable environments; policymakers or employees of professional organisations that support people with stroke; or, organisations that have experience in providing community-based programs. Participants from both groups were adults (≥ 18 years) living in Australia, able to provide informed consent and participate in workshops or interviews conducted in English either in person or via teleconferencing.

Participants were recruited via flyers and advertisements shared through social media, organisations (e.g., Stroke Foundation) and professional networks of the research team. We aimed to recruit 10-20 participants with lived experience of stroke and 5-15 professionals to participate in Stage 2 and for most of those participants to return in Stage 4 (5-15 people from each group). These numbers are consistent with previous stroke co-design studies. ^20, 21^

Participants were able to attend only Stage 4 if they were eligible to participate but unable to attend Stage 2 workshops. We aimed to purposively recruit a diverse sample of participants in terms of gender, age, time post-stroke and professional background to ensure that a broad range of knowledge and experience to optimise the relevance and reach of the co-designed walking program. ^27^ Participants were offered gift vouchers for participation in workshops.

### Procedure – Data Collection and Analysis

Workshops and one-on-one interviews were conducted both online (using ZOOM version 6 series) and in-person. They were facilitated by research team members experienced in co-design with backgrounds including physiotherapy and urban design research, clinical physiotherapy and lived experience of stroke. Workshops and interviews were recorded and field notes inclusive of reflexive comments were taken by two researchers (NF/PC). Workshop facilitators (NF/ER/DG/BAB/LS) iteratively analysed and then summarised the workshop findings and discussion to confirm consensus and correct interpretation. ^23^ This ensured that participant feedback was incorporated prior to moving on. Field notes were synthesised and cross-checked with recordings. Written workshop and interview summaries were generated by one researcher (PC), cross-checked for accuracy and clarity by a second researcher (NF) and provided to participants within 2 weeks. Participants had the opportunity to provide feedback over another 2-week period, which was recorded and incorporated.

#### Stage 1 Planning

Two workshops were held in February 2025 with the research team. In these workshops the research questions were defined, and the scope of the study was determined. Following Stage 1 the full protocol was developed, and ethical approval obtained.

#### Stage 2: Program Content Development

This stage consisted of workshops and one-on-one interviews with study participants. Separate workshops were held for lived experience and professional participants. This stage aimed to:

1. Understand the attitudes of stroke survivors, their families and carers with respect to participating in community walking groups and socialising with peers.
2. Identify the important principles of the Supportive Strides program.
3. Understand barriers and facilitators and practical or logistical considerations.

Each participant was sent workshop information and questions beforehand. This allowed time to reflect on responses prior to participation. During the workshops seeding questions were posed to encourage discussion and identification of key principles for the Supportive Strides program (see appendix 2: Workshop Seeding Questions).

#### Stage 3: Draft Program Development

Stage 3 consisted of two workshops undertaken by the research team with the aim of developing a working draft of the Supportive Strides program. In the first workshop overall findings from Stage 2 were summarised and presented to the team. During the second workshop the research team were tasked with identifying priority elements of the Supportive Strides program using a Nominal Group Technique methodology to reach consensus. ^25^ Padlet (live version 2025) was used as an online platform to facilitate idea sharing and consensus building. The five steps used in Nominal Group Technique included introduction and briefing, silent idea generation (individual level), idea sharing (sharing of ideas without discussion), group discussion (a chance to clarify ideas and discuss together) and ranking (individual ranking based on perceived importance). ^25^ Multiple rounds of ranking were permitted to guide shared decision-making of the key elements to include in the program. Following the second workshop consensus decisions were shared with the research for feedback on the consensus items. The draft Supportive Strides program was then developed.

#### Stage 4: Review and Adaptation

This stage involved mixed participant workshops and one-on-one interviews. The aim of this stage was to refine the Supportive Strides program to optimise its acceptability and potential for future implementation. Each participant was sent a copy of the draft Supportive Strides program prior to workshops. Broad questions were posed asking participants for feedback on how the program could work and what could be improved (appendix 2).

Following the Stage 4 workshops, findings were summarised and sent to the research team with the opportunity for a final round of feedback to ensure that consensus on the final program was achieved.

## Results

### Stage 1 Planning and Team Assembly

The overarching research question defined by the research team was “*What are the key principles and elements of a community walking program for people after stroke that includes peer support and socialisation?*” Sub-questions were devised to direct workshop seeding questions and understand what is important and feasible, how to incorporate a social element and how technology can be used to support the program. Further questions centred around logistical considerations to factor in, such as how and where the program could be delivered and accessed and how to make the group sustainable. The project aim was determined: to co-design a community-based walking program with embedded socialisation and peer support for people post-stroke (Supportive Strides).

### Stage 2: Program Content Development

Four workshops and one individual interview were conducted in Stage 2 with a total of 25 participants from three states across Australia (table 1). Two workshops (one in-person, one online) and one online interview were held with 12 stroke survivors and two carers. Two online workshops were held with 11 professional participants (three from professional organisations that support people with chronic disease, two urban designers, six health professionals). Some participants although primarily enrolled as a stroke survivor, carer or representative from an organisation, also identified as a health professional. As such, we had representation from physiotherapy, exercise physiology, occupational therapy and psychology (table 1).

**Table 1:** Participant Characteristics.

| Characteristic | Stroke Survivors<br>(n=12) | Carers (n=2) | Professionals (n=12) |
| --- | --- | --- | --- |
| Gender, *n female (%)* | 6 (50) | 2 (100) | 11 (92) |
| Age (years), *n (%)* |  |  |  |
| • 18-40 | 4 (33) | 1 (50) | 7 (58) |
| • 40-60 | 4 (33) | 0 (0) | 5 (42) |
| • >60 | 4 (33) | 1 (50) | 0 (0) |
| Metropolitan/rural *n (%)* | 11 (92) / 1 (8) | 2 (100) / 0 (0) | 10 (83) / 2 (17) |
| Time post-stroke, *n (%)* |  | N/A | N/A |
| • <2 years | 3 (25) |  |  |
| • 2-5 years | 4 (33) |  |  |
| • >5 years | 5 (42) |  |  |
| Self-reported walking limitations *n (%)* | 9 (75) | 2 (100) *\*person they cared for* | N/A |
| Aphasia *n (%)* | 3 (25) | N/A | N/A |
| Profession, *n (%)* | N/A | N/A |  |
| • Exercise Physiologist |  |  | 2 |
| • Occupational Therapist |  |  | 1 |
| • Physiotherapist |  |  | 6 |
| • Urban Designer |  |  | 2 |
| • Professional Organisation |  |  | 3 |
| • Psychologist |  |  | 1 |
| Years of clinical experience (years) | N/A | N/A |  |
| • < 5 years |  |  | 3 (30) |
| • 5-10 years |  |  | 0 (0) |
| • >10 years |  |  | 7 (70) |

*Footnote: Participants could be primarily enrolled as a stroke survivor, carer or member of a professional organisation, and also be a health professional.*
*19 participants were involved in both Stages 2 and 4, 6 participants were involved in only Stage 2, one participant was only involved in Stage 4.*

There was a positive attitude from all participants about the concept of a community walking program for people after stroke that includes peer support and socialisation. The following principles were seen as important for inclusion in the program

#### i. Social Support

All participants concurred that including a social element in the program would be crucial to its success. This sentiment was highlighted in participant reflections:

*“People join for the physical benefits but stay for the social benefits.”*

*Stroke survivor, male aged >60*

*“If I was joining a walking group, most important is the social aspect.”*

*Stroke survivor, male aged 40-60*

Specifically, building community, a sense of belonging and lasting social bonds were highlighted as crucial for program sustainability.

*“…a program that creates a sense of meaning and belonging for the participant”*

*Health professional, aged 18-40*

A welcoming environment where conversation was encouraged and supported was viewed as critical to success. The use of ice breakers and group warmups were proposed to facilitate initial connections. Many stroke survivors spoke of the desire to be in a group with other stroke survivors who understand their experiences.

*“…having an understanding that everyone’s been through something similar builds confidence”*

*Stroke survivor, female aged 18-40*

Including an opportunity for a coffee catch up with the opportunity for conversation alongside a walk was popular among all participants. Possible additional elements such as group chats, online forums or buddy catch ups were proposed to enhance peer support.

Group size was raised by some lived experience participants as an important consideration, particularly for those with communication difficulties. Smaller groups were proposed by some (i.e., five participants) to enable easier casual conversations.

#### ii. Inclusive Environment

Accessibility and safety of walking routes were considered essential by all participants. Factors such as flat, accessible walking paths with resting points, amenities and parking close by were proposed as important. Many participants spoke about sunshine and greenery – walking outdoors in parks, nature and gardens. Urban design participants highlighted the significance of engaging all senses to encourage a sense of delight while walking. Further insights from urban designers were the inclusion of trails that had visually appealing elements, colour, and elements of surprise to eliminate tediousness.

Offering detailed information about the walking route was raised as important for both clinician referrers in terms of safety and preparing and practising the required elements, and stroke survivors to assist with confidence-building and preparation. The ability to cater for all levels of ability was discussed. Flexibility was proposed as an important element to enable people of different abilities to join, for example walking at their own pace and meeting back at a set point.

*“It is important to challenge people appropriately”*.

*Health professional, aged 40-60*

Indoor or sheltered options were considered pertinent to offer alternative locations in instances of rain or extreme heat, to maintain consistency in walking frequency. Shopping centres were proposed as an option due to their accessibility and amenities. Whilst the idea of enabling broad inclusivity in terms of walking routes and environments was seen as positive by many, a cautionary note was raised:

*“When you design for universal access, you design for nobody – you lose the specificity of the design intent.”*

*Urban designer, aged 40-60*

#### iii. Motivational Support

Providing support for sustaining motivation to attend the walking group was highlighted as important for ongoing participation by participants. The group should feel meaningful, fun and rewarding and shouldn’t be just seen as an obligation.

*“Participation needs to come from motivation rather than obligation”*

*Urban designer, aged 18-40*

Goals were highlighted as a potential motivating factor and having assistance with goal setting was thought to be a facilitator to participation.

*“Achieving your goals can be very motivating”*

*Stroke survivor, female aged >60*

Tracking and celebrating or recognising achievements was proposed as a method to support motivation. Simple measures to track progress such as distance walked, time taken and number of walks completed were highlighted as motivating by people with stroke.

*“I think what’s worked for my clients is keeping record of their own walking progress…. so they can reflect on their previous walking times or distances. They can see how much they’ve progressed…. which keeps them motivated.”*

*Health professional, aged 18-40*

Acknowledging individual (non-competitive) milestones and group milestones (such as number of group walks completed) were also proposed to enhance motivation. Celebrating events like birthdays and end-of-year were seen as valuable for building a sense of community and motivation to keep attending.

Accountability in the form of having someone to regularly check in was also suggested to support motivation. This did not necessarily need to be a face-to-face contact – it could be a phone call or text message to check in. These accountability “check-ins” were viewed as important for encouraging people to not only go for a walk on “group days,” but to also walk independently at other times of the week. Further, it was emphasised that motivation could be provided by not only a group leader, but also peers or “buddies” in the group.

*“You need someone to say hey – how did you go today? That’s great to hear! It’s that encouragement to get out and do it – that accountability.”*

*Stroke survivor, male aged 40-60*

Building a waking group into a person’s routine was viewed as critical to sustainability, so elements like having a consistent time, day and location each week were seen as a desirable. The challenge of fitting into busy work and personal lives was acknowledged by many, further reinforcing the need for motivational support.

#### iv. Co-ordination and administrative support

It was seen as essential to have a group coordinator to oversee the program and provide administrative support, particularly for the group’s sustainability. Key responsibilities may include welcoming new participants, sharing routes and schedule of walks, sending reminders, moderating the group chat, conducting accountability check-ins, co-ordinating and training buddies, setting goals, tracking milestone progress and organising celebrations. Further to this it was proposed that the coordinator could encourage conversation skills and confidence in social settings to ensure everyone is included.

*“Because you always need a leader to organise who, when and where”*

*Stroke survivor, male aged 40-60*

*“Perhaps it needs, like, some sort of goal setting and a plan for each person” Carer, female*

Another important role for the coordinator was to ensure safety for stroke survivors through a clear registration process. This may involve ensuring that everyone has carefully read and accepted relevant ‘terms and conditions’ for participation. For some at higher risk (e.g., of falls) it may be recommended that a medical clearance is obtained prior to participation. Some lived experience participants thought it essential that the group be run by a health professional for safety and to answer any questions that arise, whereas others thought referral into the group by a health professional would be sufficient.

*“It would possibly build confidence – if you had a physio or OT there, they’d have more of an idea of what to do if you had a problem.”*

*Stroke survivor, male aged 40-60*

Further ideas from the workshops included having a website to promote the group and house all relevant information, including support workers and carers, and engaging corporate support to enable ongoing funding for a website and co-ordination.

Barriers to participation were identified and potential mitigating solutions were proposed; these are highlighted in table 2. Barriers centred around mobility, safety, information, logistics and sustainability.

**Table 2:**
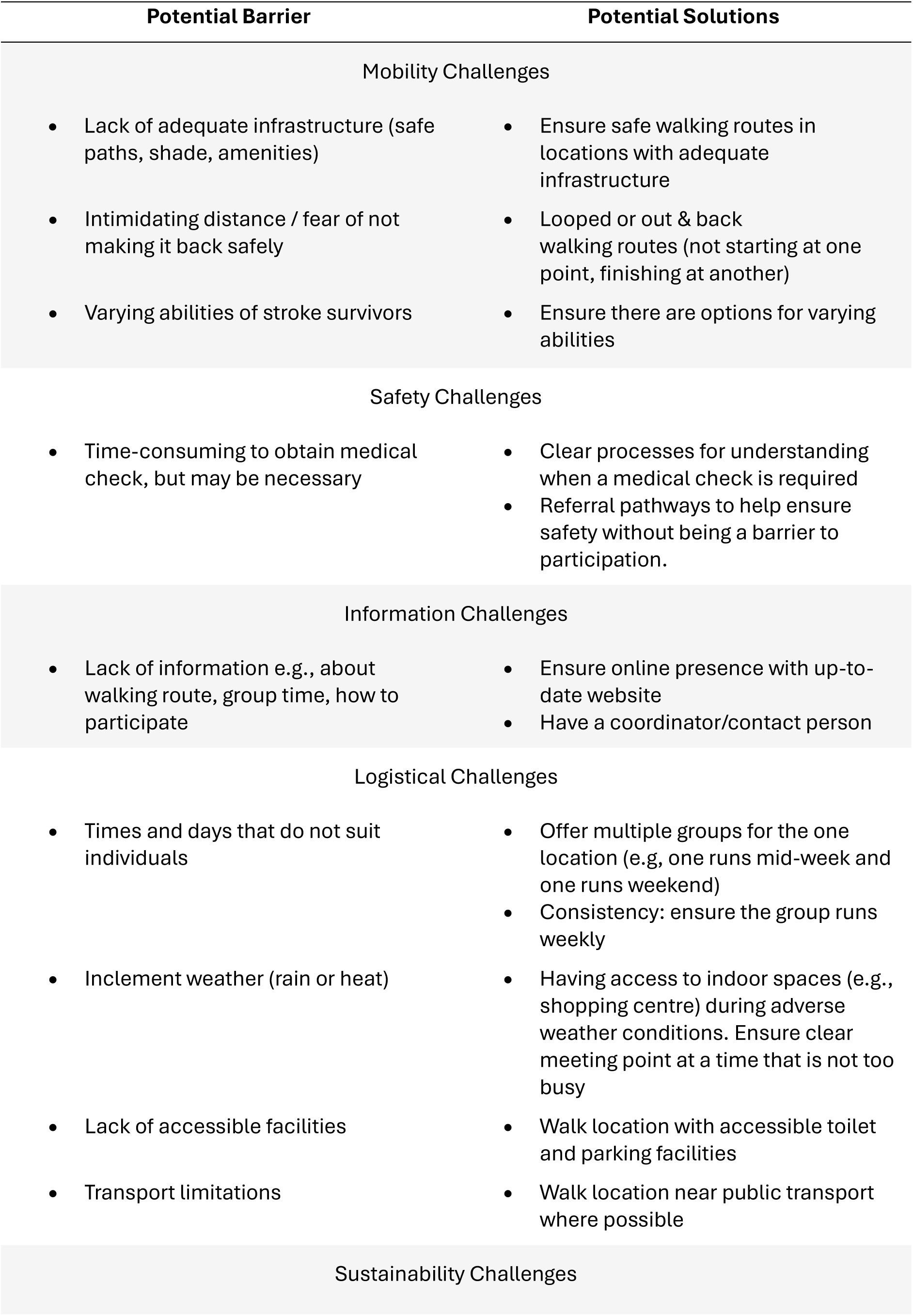
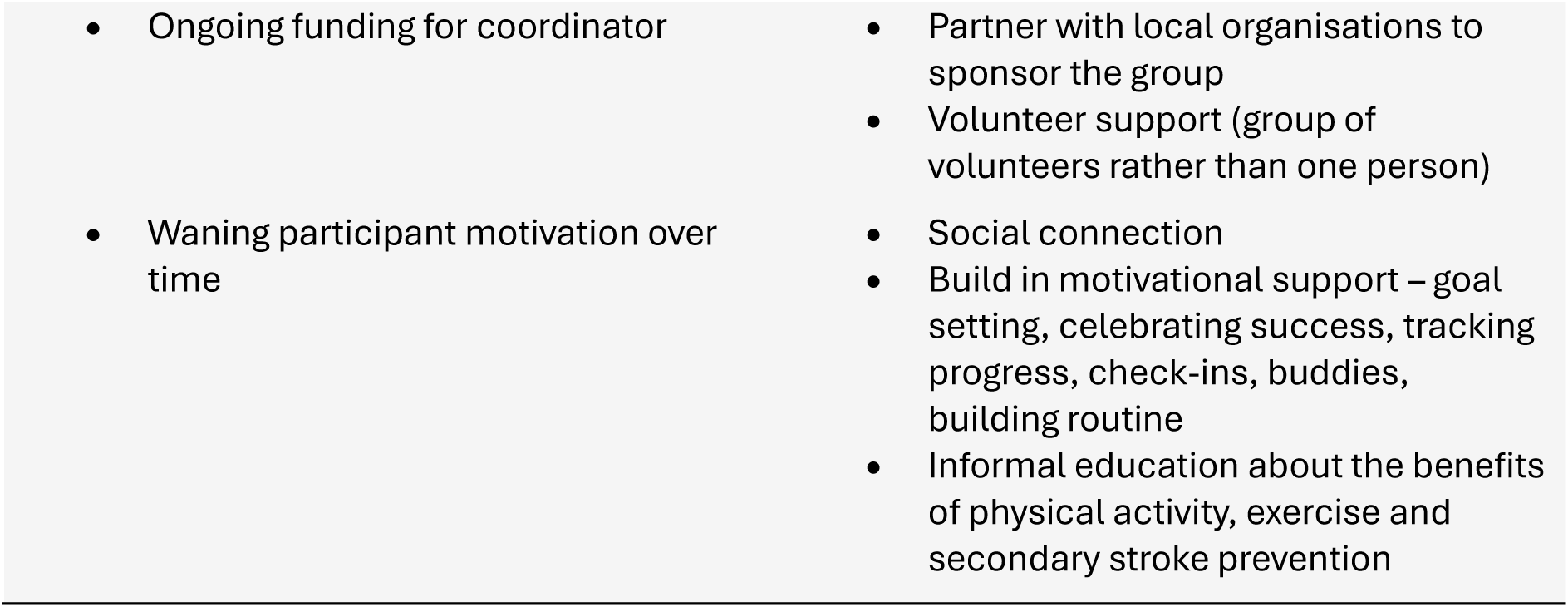
Potential Barriers to Participation in the Supportive Strides Program and Potential Solutions.

### Stage 3: Draft Program Development

The research team determined fourteen program elements through the Nominal Group Technique process. These program elements are outlined in table 3 and mapped to program principles.

**Table 3:**
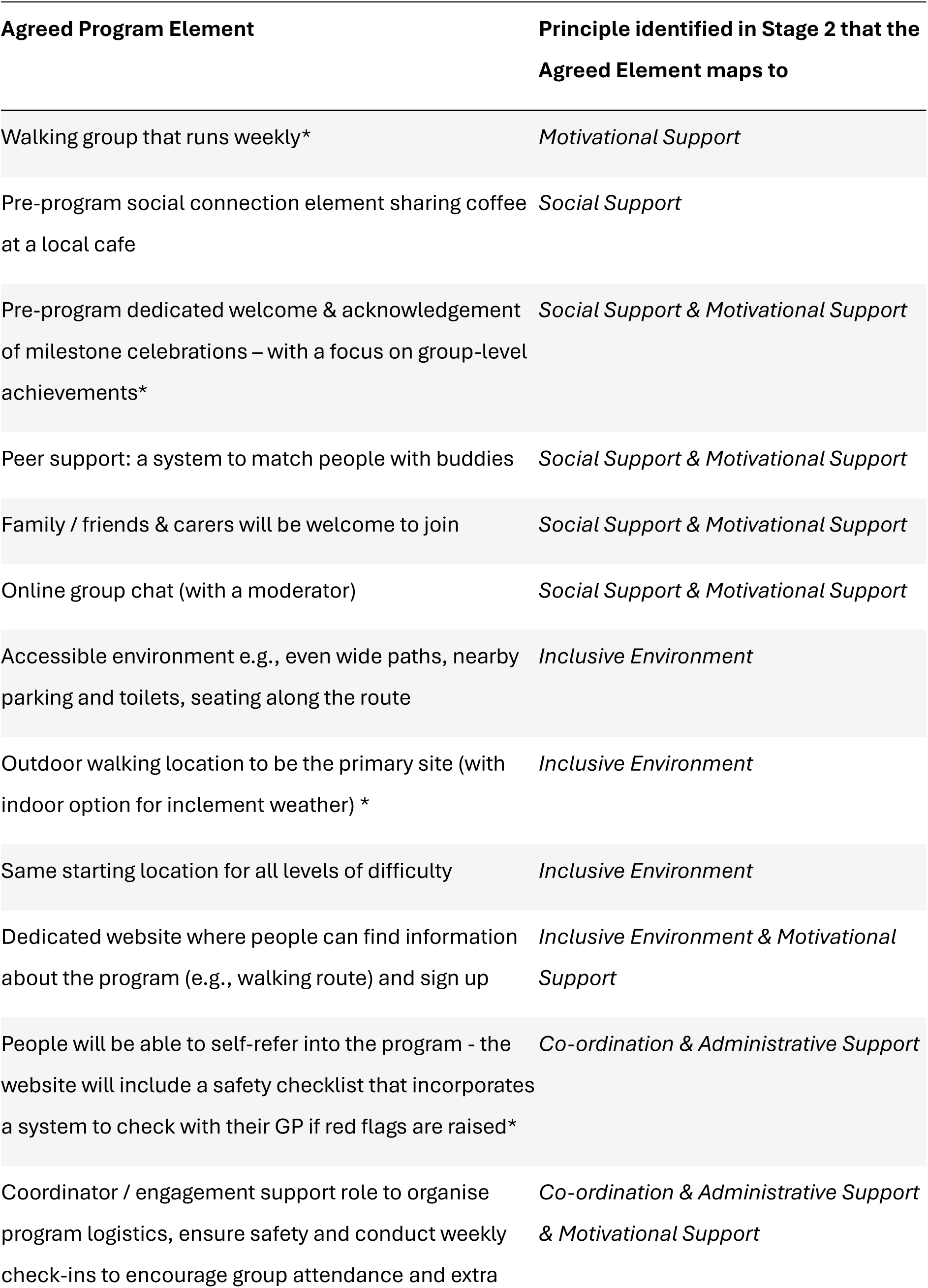
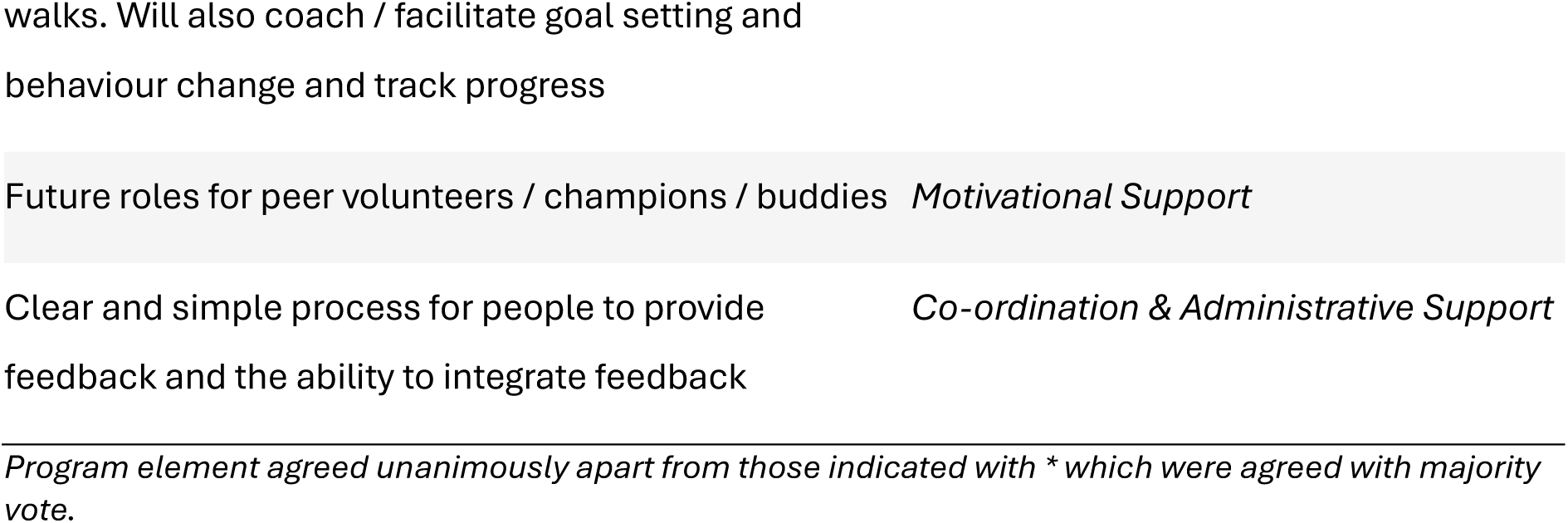
Supportive Strides Agreed Program Elements.

The walk location was a point of discussion – the desire for an outdoor location was clear among stage 2 participants however the need for an indoor location to account for adverse weather was noted. This highlighted the need for a group chat where any decisions regarding location could be communicated.

After extensive discussion it was decided that the co-ordinator should have a health professional qualification. This was thought necessary to enable discussion of safety and possible medical clearance. Further, the role will include some aspects of coaching, facilitating goal setting and behaviour change in the check-in sessions, as well as facilitating group conversation skills.

Consensus was not initially reached on whether the program should have a formal education element. However, the lived experience researchers provided further reasons for not including an education component and reasoned that there are many existing platforms that have stroke resources and information widely available for consumers (e.g., https://enableme.org.au/).

Following these insights, the group consensus was not to include education.

Some logistical aspects of the program were discussed such as whether the group should run on weekends vs weekdays, the size of the groups and running the program in regional vs metropolitan areas. Many of these decisions would be made at a local level and be specific to each group. It was also agreed that beyond any initial grant funding, partnerships and sponsors would be critical for sustainability of the Supportive Strides program.

### Stage 4: Review and Adaptation

Three workshops and one one-on-one interview were conducted online in Stage 4. Workshops were held with a mix of lived experience and professional participants. Nineteen participants from Stage 2 (8 stroke survivors, 1 carer, 8 professionals) and one professional participant who was unable to attend Stage 2 contributed to Stage 4.

Participants thought the Supportive Strides program would be beneficial for people after stroke. They particularly valued the social element of the program, specifically the opportunity to build connections, reduce isolation and improve motivation. Strong support was evident for the pre-walk coffee catch-up and acknowledgement that walking as a group facilitates accountability and consistency. There was consensus that the outdoor environment was preferred, but a backup wet weather option was important. A website with walking route information that could be viewed prior to attendance was seen as necessary.

A buddy system was considered important for peer support and discussion centered around how buddies should be matched. The possibility for a one-on-one orientation with buddies prior to the first walking group session was raised as a method to build confidence. Fear of ‘starting something new’ was thought to be a significant barrier and methods to provide support for this were highlighted as critical.

Some challenges were acknowledged – particularly timing as limiting attendance to the same day and time each week will not suit everyone. Many lived experience participants noted that early mornings are not suitable. The registration process/safety checklist was also raised as a potential barrier to participation, as overly complex registration processes can be a deterrent.

Additions were made to specific program elements following Stage 4 workshops. These included: matching buddies by age and ability level; including more walking route information online (e.g., videos and maps that highlight seating and other accessibility information); tracking specific metrics (e.g., distance walked, step count) and ensuring that the safety checklist has no more than 10 questions and includes confidence, exercise concerns and support required.

## Discussion

In this study we co-designed the Supportive Strides community-based walking program for people with stroke. Researchers with lived experience were key decision-makers throughout the process and their input ensures the Supportive Strides program is relevant to people with stroke. This program uniquely adds socialisation and peer support to a community walking program. Such additions were viewed as critical elements for program motivation and sustainability. Inclusive environmental features were raised as necessary for safety and participant enjoyment, and specific elements around tracking and celebrating progress, and coaching support were viewed as important for motivation.

Inclusion of a social connection element was considered as a critical active ingredient of the Supportive Strides intervention, and we propose that it will be a key driver of participant motivation, successful outcome and program sustainability. Many post-stroke and physical activity programs only include a physical component, and this may be one reason why physical activity habits are not sustained after a program ends. ^4^ Including a social element has been shown to improve motivation for physical activity in healthy older populations when evaluating the social outcomes of group outdoor walking initiatives. ^9–11, 14^ With the addition of other motivational support elements (e.g., buddy’s, acknowledgement of milestones) we anticipate the Supportive Strides program will improve physical activity outcomes for people with stroke. Careful consideration around all program elements is required for future program sustainability. The Program Sustainability Assessment Tool, developed to assess public health programs’ capacity for sustainability over time, may enable successful implementation of Supportive Strides. The Tool assesses eight domains critical for program sustainability and can be used to guide program development and key considerations from the outset. ^28, 29^ Items from the Program Sustainability Assessment Tool, including having strong champions and adequate staffing or coordination support, have already been considered for Supportive Strides. Areas important to establish in the future will be sources of ongoing funding and partnerships.

The environmental and behaviour change elements of our co-designed Supportive Strides program align with the policy action areas of the recently published World Health Organisation toolkit of policy options for promoting walking and cycling. ^30^ Highlights of their policy action areas include providing safe and connected walking networks with integrated green spaces that are designed to be inclusive by ensuring safe, accessible and attractive environments for all abilities. ^30^ The participants in our study articulated that accessible green spaces are necessary not only for safety and confidence, but also for enhancing motivation and making walking a priority. By giving stroke survivors the opportunity, confidence and motivation to participate in a walking program, it may facilitate them to use walking as a means of active transport which will benefit not only their health but also the environment. These elements align with the COM-B framework^31^ which supports the premise that Supportive Strides may be effective at changing long-term physical activity behaviours. The Supportive Strides program also includes elements of the WHO’s key advocacy strategies for walking on a small scale by appointing champions and engaging local communities. ^30^ This demonstrates the future potential of such a program.

Co-design is thought to be vital for complex healthcare intervention design. ^17^ This research is positioned at two different sections of the recently developed Co-Creation Rainbow Framework. ^32^ Inclusion of stroke survivors as participants in co-design places this research in the stimulating and collaborating sections of the continuum, while the inclusion of two people with lived experience of stroke on our research team places the research at the empowering or collective decision-making section of the continuum. ^32^ Our study demonstrates partnership, an element deemed imperative for true co-design and particularly highlighted in the IKT approach. ^19, 33^ Our lived experience researchers were key in the decision-making processes and were involved in tasks such as reviewing patient information and consent forms, tailoring questions for workshops, facilitating workshops, consensus building, manuscript review and authorship. ^18, 34^ There are clear examples of their impact on the end-result. When facilitating workshops, they were able to draw rich responses from participants, possibly due to participants being willing to open up more to a person with a shared experience of stroke. Our lived experience researchers also enabled us to reach consensus about not including an education component in Supportive Strides. Without their input, we may have included an unnecessary program element, which will have led to resource waste. Our co-design study adds to the body of literature where inclusion of lived experience researchers with the right supports and processes^22, 34, 35^ has resulted in successful stroke recovery intervention-development. ^20–23, 35–37^

While this study adhered strongly to co-design principles and included insights from a broad range of participants and researchers, including design experts who do not traditionally contribute to healthcare intervention development, there were some limitations. These include the modest sample size and time constraints which limited flexibility in the timing of workshops. ^18^ The limited flexibility may have reduced participation and resulted in a potentially biased cohort.

To conclude, the Supportive Strides community-based walking program was co-designed with the aims of improving physical activity, social networks, confidence, wellbeing and health after stroke. This study provides a clear direction for future research through the testing of the Supportive Strides program in real-word environments. Additional work may include review and / or production of a suitable walkability framework to assess local walking environments.

## Data Availability

All data produced in the present study are available upon reasonable request to the authors

## Acknowledgements

The authors would like to acknowledge the stroke survivors, carers and professionals who participated in this co-design study.

## CRediT Roles

Conceptualisation: NAF, ER, PC, BB, LS, DG, AM, IMB, CHM, KSH

Methodology: NAF, ER, DG

Software: NAF, PC

Validation: NAF, ER, PC

Formal Analysis: NAF, ER, PC

Investigation: NAF, ER, PC, BB, LS, DG, AM

Resources: NAF, ER, PC, BB, LS, AM, IMB, CHM, KSH

Data Curation: NAF, PC

Writing-Original Draft: NAF

Writing – Review & Editing: NAF, ER, PC, BB, LS, DG, AM, IMB, CHM, KSH

Supervision: NAF

Project administration: NAF, PC

Funding acquisition: NAF, ER, BB, LS, DG, AM, IMB, CHM, KSH

