## Appendices for "Supportive Strides: Co-designing Community-Based Walking Programs with Embedded Socialisation and Peer Support for People after Stroke"

#### Appendix 1 GRIPP2 Reporting Template

| Section and Topic | Item | Reported on Page No. |
|---|---|---|
| **Section 1: Abstract** | | |
| 1a. Aim | Report the aim of the study | Abstract p1 |
| 1b. Methods | Describe the methods used by which the patients and the public were involved | Abstract p1 |
| 1c. Results | Report the impact and the outcomes of PPI in the study | Abstract p1 |
| 1d. Conclusions | Summarise the main conclusions of the study | Abstract p1 |
| 1e. Keywords | Include PPI, “patient and public involvement,” or alternative as keywords | Keywords (co-design) p1 |
| **Section 2: Background** | | |
| 2a. Definition | Report the definition of PPI used in the study and how it links to comparable studies | Introduction p4 & Methods – study design p4 |
| 2b. Theoretical underpinnings | Report the theoretical rationale and any theoretical influences relating to PPI in the study | Introduction p3-4 & Methods – study design p4 |
| 2c. Concepts and theory development | Report any conceptual models or influences used in the study | Introduction p3-4 & Methods – study design p4 |
| **Section 3: Aims** | | |
| 3. Aim | Report the aim of the study | Introduction p4 |
| **Section 4: Methods** | | |
| 4a. Design | Provide a clear description of methods by which patients and the public were involved | Methods p4-7 |
| 4b. People involved | Provide a description of patients, carers and the public involved with the PPI activity in the study | Methods p4-5 |
| 4c. Stages of involvement | Report on how PPI is used in different stages of the study | Methods p4-7 |
| 4d. Level or nature of involvement | Report on the level or nature of PPI used at various stages of the study | Methods p4-7 |
| **Section 5: Capture or Measurement of PPI Impact** | | |

| | | |
|---|---|---|
| 5a. Qualitative evidence of impact | If applicable, report the methods used to qualitatively explore the impact of PPI in the study | Methods p4-6 |
| 5b. Quantitative evidence of impact | If applicable, report the methods used to quantitatively measure or assess the impact of PPI | Methods p4-6 |
| 5c. Robustness of measure | If applicable, report the rigour of the method used to capture or measure the impact of PPI | Methods p4-6 |
| **Section 6: Economic Assessment** | | |
| 6. Economic assessment | If applicable, report the method used for an economic assessment of PPI | N/A |
| **Section 7: Study Results** | | |
| 7a. Outcomes of PPI | Report the results of PPI in the study, including both positive and negative outcomes | Results p7-13 |
| 7b. Impacts of PPI | Report the positive and negative impacts that PPI has had on the research, the individuals involved (including patients and researchers) and wider impact | Results p8-13 |
| 7c. Context of PPI | Report the influence of any contextual factors that enabled or hindered the process or impact of PPI | Results p8-13 |
| 7d. Process of PPI | Report the influence of any process factors, that enabled or hindered the impact of PPI | Discussion, limitations p15 |
| 7ei. Theory development | Report any conceptual or theoretical development in PPI that have emerged | Discussion p14-15 |
| 7eii. Theory development | Report testing of any theoretical methods if any | N/A |
| 7f. Measurement | If applicable, report all aspects of instrument development and testing (e.g., validity, reliability, feasibility, acceptability, responsiveness, interpretability, appropriateness, precision) | N/A |
| 7g. Economic assessment | Report any information on the costs or benefit of PPI | N/A |
| **Section 8: Discussion and Conclusions** | | |
| 8a. Outcomes | Comment on how PPI influenced the study overall. Describe positive and negative effects | Discussion p13-15 |
| 8b. Impacts | Comment on the different impacts of PPI identified in this study and how they contribute to new knowledge | Discussion p13-15 |
| 8c. Definition | Comment on the definition of PPI used (reported in the background section) and whether or not you would suggest any changes | Discussion, p13-15 |
| 8d. Theoretical underpinnings | Comment on any way your study adds to the theoretical development of PPI | Discussion, p15 |
| 8e. Context | Comment on how contextual factors influenced PPI in the study | Discussion, p13-15 |

| | | |
|---|---|---|
| 8f. Process | Comment on how process factors influenced PPI in the study | Discussion, p13-15 |
| 8g. Measurement and capture of PPI impact | If applicable, comment on how well PPI was evaluated or measured in this study | N/A |
| 8h. Economic assessment | If applicable, discuss any aspects of the economic cost or benefit of PPI, particularly any suggestions for future economic modelling | N/A |
| 8i. Reflections / critical perspective | Comment critically on the study, reflecting on the things that went well and those that did not, so that others can learn from this study | Discussion, p13-15 |

**PPI = patient and public involvement*

**Appendix 2 Workshop Seeding Questions**

### STAGE 2 WORKSHOP SEEDING QUESTIONS

**Italics are just prompts for research team/facilitator)**

1. ***LIVED EXPERIENCE PARTICIPANTS***

START WITH THESE BROAD QUESTIONS:

- What would a great day look like if you were participating in a walking group or program?
- How would you see a walking group or program fitting into your life long term?

*These questions might address things like time of day, daily behaviours, places they are familiar with etc*

*THEN MORE TAILORED QUESTIONS, SUCH AS:*

- How could we include a social element in a walking program?

*e.g., coffee catch ups*

Peer support will be a feature of our program,

- Who do you see as your peers? How might they support you in this program?

*e.g., face to face, online etc*

- How could you see a buddy-system working for a program like this? How could we "match" buddies?
- How often would you like to participate in a walking program?
- Where would you like to walk?

*e.g.,outdoors or indoors eg at a shopping centre, consistent location?*

- What are important considerations?
- Would you like to be able to talk to someone about why you are doing a walking program, what you want to achieve and how to keep going with a program in the long term?

*e.g., talk to someone face to face, phone or just an app or even AI*

- Would you like this program to be incorporated into an existing initiative
    - *(eg Parkrun, HF, iRebound)*
- What would help (motivate) you to take part in and stick to a walking program like Supportive Strides?

- Are rewards or incentives important?
- What are your barriers (roadblocks) to taking part in a walking program like Supportive Strides?

2. ***PROFESSIONAL PARTICIPANTS***

BROAD QUESTIONS:

- *What does the ideal, sustainable, community walking group for people who have had a stroke look like?*

*THEN MORE TAILORED QUESTIONS:*

- What has worked for you with bringing people together to be physically active?
- What do you think are key elements to successful programs?
- Are there any specific urban design elements we should be considering, especially considering the varying physical abilities of stroke survivors?
- What would you avoid doing – ie what hasn't worked in the past?
- What are the potential barriers to a program like this?
- Do you have any words of wisdom for us?

**STAGE 4 MIXED WORKSHOP SEEDING QUESTIONS**

- Which aspects of the proposed program will work for you?
- Which aspects of the proposed program won't work for you?
- Any suggestions on how we could improve the program to make it work better for you?
